# Assessing Cellular and Transcriptional diversity of IIeal Mucosa amongst Treatment Naïve and Treated Crohn’s disease

**DOI:** 10.1101/2022.05.19.22275263

**Authors:** Sushma Chowdary Maddipatla, Vasantha L Kolachala, Suresh Venkateswaran, Anne F Dodd, Ranjit Singh Pelia, Duke Geem, Hong Yin, Yutong Sun, Congmin Xu, Angela Mo, Astrid Kosters, Junkai Yang, Jason D. Matthews, Eliver Ghosn, Subra Kugathasan, Peng Qiu

## Abstract

**Background and Aims:** Crohn’s disease is a life-long disease characterized by chronic inflammation of the gastrointestinal tract. Defining the cellular and transcriptional composition of the mucosa at different stages of disease progression is needed for personalized therapy in Crohn’s.

**Methods:** Ileal biopsies were obtained from (i) controls (n=6), (ii) treatment-naïve (n=7), and (iii) established (n=14) Crohn’s patients along with remission (n=3) and refractory (n=11) treatment groups. The biopsies were processed using 10x Genomics single cell 5’ yielded 139,906 cells. Gene expression count matrices of all samples were analyzed by reciprocal principal component integration, followed by clustering analysis. Manual annotations of the clusters were performed using canonical gene markers. Cell-type proportions, differential expression analysis and gene ontology enrichment were carried out for each cell type.

**Results:** We identified 3 cellular compartments with 9 epithelial, 1 stromal, and 5 immune cell subtypes. We observed differences in the cellular composition between ctrl, treatment-naive and established groups, with the significant changes in the epithelial subtypes of the treatment-naive patients, including microfold, tuft, goblet, enterocytes and BEST4+ cells. Surprisingly, fewer changes in the composition of the immune compartment were observed, however gene expression in the epithelial and immune compartment was different between Crohn’s phenotypes indicating changes in cellular activity.

**Conclusions:** Our study identified cellular and transcriptional signatures associated with treatment-naive that collectively points to dysfunction of the intestinal barrier with an increase in inflammatory cellular activity. Our analysis also highlights the heterogeneity amongst patients within the same disease phenotype, shinning new light on personalized treatment responses and strategies.

## Introduction

Crohn’s disease (CD) is characterized by chronic inflammation of the gastrointestinal tract^1^, particularly the ileal or ileocecal locations^2, 3^, but distinct from ulcerative colitis (UC), which shows clinical overlap with colonic CD^4^. The incidence of CD in North America is roughly 20 cases per 100,000 ^5,6^, but reaching upwards of 200 per 100,000 in some regions of the world. The exact cause of CD is unknown but is thought to be the result of a complex and multifactorial etiology with interplay between lifestyle, genetic pre-disposition, epigenetics, epithelial barrier disruption, and unbridled immune response to intestinal triggers^7,8,9^. The disease onset can be at any age, but most of the cases are diagnosed in the pediatric population with the peak between 10-25 years old^10^. Although the clinical course of CD is characterized by relapses and remission, once the disease begins, it typically is a life-long illness. Clearly, CD has a genetic component as several GWAS and follow-up studies demonstrated risk associated loci^6, 11,10^, yet most of the 200 or so IBD risk loci fall within non-coding regions, suggesting more epigenetic and molecular characterization needs to be performed. Since the broadly used therapies like corticosteroids and immunomodulatory drugs are ineffective in controlling the disease, recently developed biologicals including anti-TNF, anti-adhesion molecules and anti-IL12/23 have become frontline targeted therapies for CD^12^. Although the recent cumulative data suggest these biologic therapies are better than conventional therapies, they are only effective in about 30-50% of the patients. Unfortunately, biologic treated patients, particularly those on anti-TNF, have an increased risk for opportunistic infections and malignancies such as lymphomas and skin carcinomas^13^. Thus, new and improved targeted therapies in CD are an unmet need, and expansion of available treatment options requires a shift towards a precision medicine approach. Furthermore, primary and secondary non-response (refractory) to biologics is increasingly common but understanding the mechanism for non-response requires more understanding of the cellular and transcriptional variability across different CD subjects. There has been limited success from clinical and biological phenotyping in identifying novel biomarkers as companion diagnostics to guide precision CD treatment.

To date, several bulk biopsy transcriptomic studies have been performed for IBD, however, those studies were limited to using whole tissue for analysis of phenotypic associations and pathways. Therefore, recent studies have aimed at enabling precision medicine in IBD by using new technologies like single-cell RNA sequencing (scRNA-seq) and mass cytometry (CyTOF) to predict therapy response and disease activity^9, 14, 15^. One recent study aimed to identify pathogenic cellular modules that were weakly, although significantly, associated with resistance to a n t i - TNF therapy^9^. The presence of these immune/ m a c r ophage cell modules suggests a spatial and functional relationship between underlying cell types. Since those studies used surgically resected tissue that is the end-stage of a chronic inflammatory process, it is vital that colonoscopically retrieved mucosal biopsies are used to study the different locations, stages, and course of CD. Application of a high multi-dimensional scRNA-seq approach provides unprecedented resolution of cell lineages to characterize the transcriptomic heterogeneity of cell types and their functional states in human tissues^16^. A deeper understanding in cellular heterogeneity and specificity at the single-cell level can be achieved if the mucosa is studied at diagnosis when patients are treatment-naïve and may provide additional insights for ileum specific inflammation during early CD when compared to established CD patients.

Thus, in this study, we have used scRNA-seq of colonoscopically retrieved terminal ileal mucosal biopsies from 27 subjects across three different phenotypes, including non-IBD controls (ctrl), treatment-naïve (TN_CD) CD, and established (Es_CD) CD. Es_CD allowed us to study the mucosa in those who had healing (remission) and in those that did not (refractory). Our analysis revealed distinct cellular and transcriptional complexities within the ileal mucosa of TN_CD patients, allowing identification of both cell types and cell-type-specific genes that exhibited changes across different patient phenotypes, and further providing a glimpse into mucosal differences at the cellular and transcriptional level between two treatment response groups, remission, and refractory in Es_CD, all of which can be used to formulate new hypotheses towards personalized treatment strategies for different stages of CD.

## Methods

### Study Population

Terminal ileal biopsy samples were collected from 27 patients who were undergoing clinically indicated colonoscopy at the Division of Pediatric Gastroenterology at Children’s Healthcare of Atlanta and Emory University (Georgia, USA). Written informed consent was obtained before sample collection. The study group includes controls (ctrl, n=6), treatment-naïve CD (patients had not yet received treatment at the time of diagnosis; TN_CD, n=7) and established CD patients (patients were diagnosed earlier and received treatment for at least one year; Es_CD, n=14). Endoscopic^17^ (SESCD in CD) and histological activity (presence or absence of histological inflammation) were documented for all the subjects. Depending on a patients’ response to the treatment, the Es_CD group was divided into remission (who responded to treatment with evidence of mucosal healing; n=3) and refractory (who did not respond and had active inflammation with colonoscopy and histology; n=11) (**Supplementary Table1**).

### Tissue Processing

Once the biopsy was collected from the patient through endoscopy, it was immediately transported from the endoscopy suite to the lab in complete RPMI media with 10% FBS and fully processed within two hours of collection. The biopsies were first minced with micro-scissors on ice in HBSS buffer containing 2mM EDTA without Ca^2+^/Mg^2+^. The minced pieces were vortexed gently and allowed to settle for 2 min on ice. The supernatant containing some of the loosely associated immune cells and other single cells released by the mincing process (primarily lamina propria) were removed and the sedimented minced tissue pieces were digested on ice with *B. licheniformis* protease for 30 minutes. Samples were periodically vortexed gently and placed back on ice during enzymatic digestion. For each patient sample, the supernatant fraction was mixed with the enzyme digested fraction 1 : 1 for non-inflamed mucosa, and 1 : 4 for inflamed mucosa (minimizing overloading with immune cells) before being passed through a 40- and then 20-µm filter, washed and finally resuspended at the appropriate cell density for loading onto the 10X Chromium Controller (~1000-1200 cells/µL).

### 10x Library Preparation and Sequencing

Tissue derived single cells were loaded onto the 10X Chromium Controller targeting 10,000 cells. Single cell capture, barcoding, GEM-RT, clean-up, cDNA amplification and library construction were performed according to the manufacturers’ instructions using Chromium Single Cell A Chip Kit (cat no: PN-1000152) and Chromium 5’ Library & Gel Bead Kit (v1 chemistry; cat no: PN-1000006). Final library pools were sequenced in four batches on Illumina’s Novaseq6000 instrument at Georgia Tech Genomics Core or Yerkes Genomic Core with a targeted sequencing depth of 50,000 reads/cell.

### 10x Genomics Computational Analysis and Quantification

Cell Ranger (10x Genomics)v6.0 was used to generate gene expression count matrices for every sample, after sequence alignment to Grch38 human reference genome using STAR^18^ to obtain UMI (Unique Molecular Identifiers) counts for each gene per cell. Based on UMI counts and cell barcodes, numbers of genes and cells for each sample were estimated. Matrices for each sample were analyzed by Seurat v4.1. Since the intestine has high cell turn over, it is important to consider all the cells, including the cells that are under stress. Therefore, quality control threshold for mitochondrial percentage was not set for the analysis as intestinal epithelium undergo constant regeneration and apoptosis. In addition, we observed a small population of epithelial cells showing very low expression of immune markers and vice versa but it did not affect the overall cluster annotations.

### Clustering Analysis

The Seurat v4.1 RPCA integration^19,20, 21^ workflow was applied to cluster cells from all the samples. The workflow started with log-normalization of the data and highly variable feature selection independently for each sample. Features that are repeatedly selected as variable genes across the samples formed the integrated feature list for scaling and PCA, which were further used to find anchors that assisted the integration of data from all samples. This integration workflow corrected for batch effects in the data. Principal component analysis was performed on the integrated dataset to reduce the data into 30 dimensions (i.e., top 30 PCs). The PCA space was used to generate cell clusters and generate Uniform Manifold Approximation and Projection (UMAP) to visualize the cell clusters.

### Marker Genes and Annotations of Clusters

After cell clustering, differential gene expression analysis was performed between each cell cluster against all other cells to obtain marker genes for interpreting the cell clusters. The differential expression analysis was performed on the uncorrected data before integration, but after library size normalization with scaling factor of 10,000 and log transformation. Wilcoxon rank sum test was used to identify and rank differentially expressed genes, using Seurat “FindMarker” function. Cell clusters were annotated by comparing the marker genes derived from our dataset with publicly available databases and the marker genes published by Martin et al., ^9,^ and Wang et. al ^22^. The marker genes were manually examined by color-coding the UMAP visualization allowing for confirmation of appropriate expression patterns across cell clusters and that they agreed with the cell type annotations.

### Differential Gene Expression across CD Phenotypes

Differential gene expression analysis was performed on every cell type from each pair of CD phenotypes (TN_CD vs ctrl; TN_CD vs Es_CD; Es_CD vs ctrl), using Wilcoxon rank sum test implemented by the Seurat “FindMarker” function (min.pct=0.1, log.threshold=0.25). We used the same settings to identify differentially expressed genes between remission and refractory in Es. These analyses were performed for each cell cluster separately. Seurat’s BuildClusterTree function was used to construct phylogenetic trees that represent distances between the average cells for each epithelial (**Supplementary Figure 5A**) and immune sub-types (**Supplementary Figure 5B**).

### Ontology Enrichment Analysis

GO enrichment analysis was performed using the cell-type-specific differentially expressed genes between the CD phenotypes with p-value <0.05, using R package ‘cluster profiler’^23^. The results were viewed using cluster profiler “dotplot” function and “cowplot” packages.

## Results

### Treatment-naïve CD mucosa shows disrupted epithelial subtype proportions while B cells are decreased in established CD

First, we sought to define the differences in cellular composition of the terminal ileum in TN_CD & Es_CD in comparison to non-IBD controls, and to better understand the association of different cell types with disease phenotypes. To this end, we generated 139,906 single cell transcriptomes from ctrl and CD ileal mucosal biopsies. The scRNA-seq data went through quality control (**Supplementary Figure 1A-C)** and integration pipelines (**Supplementary Figure 1D-E**) to account for batch effects. The integrated data was clustered and visualized by UMAP (**Figure 1A**). When the UMAP visualization was stratified by patient phenotype groups (**Figure 1B**) a high degree of overlap between ctrl and CD can be observed. We performed differential gene expression between the cell clusters to identify marker genes that can be used to annotate the cell clusters with known cell types, based on known cell type signature genes from published literature^9, 22^. We were able to manually annotate 9 epithelial, 5 immune cell types and 1 stromal cell type (**Supplementary Figure 2A**). Epithelial cells were subdivided into 9 cell types with the shared expression of *EPCAM, KRT8* & *KRT18* expression in both ctrl and CD. The genes used to define each subcluster of Epithelial cells were: stem/TA/Progenitor (*OLFM4, LGR5*); TA cells (*OLFM4, MKI67* and *TOP2A*); early enterocyte (*OLFM4, LCN2* and *ADH1C*); enterocyte (*APOA4* and *FABP6*); Goblet cells (*MUC2, TFF3*, and *SPINK4*); enteroendocrine cells (EECs) (*CHGA, CHGB, PYY*, and *NTS*); paneth cells (*DEFA5, DEFA6*, and *LYZ*); BEST4+ enterocyte (*BEST4, OTOP2*, and *SPIB*)^24^; tuft cells (*SPIB, SH2D6*, and *LRMP*). For the immune compartment defined by *PTPRC* expression, 5 subtypes were defined by the following gene expression patterns: B cells (*MS4A1*); T cells (*CD3D* and *NKG7*); plasma cell (*JCHAIN, MZB1, IGHA1*, and *IGHG1*); macrophages (*S100A8, C1QA*, and *C1QB*); erythrocytes (*HBB* and *HBA1*). The stromal cells were defined by *ADAMDEC1, ACTA2*, and *COL3A1* expression. In this global analysis, at the level of cell type, with the exception of enterocytes that are significantly reduced in TN_CD (p<0.05), and B cells that are significantly reduced in Es_CD compared to ctrl^25-27^, most of the detected cell types were found in similar proportions between the three patient phenotype groups (**Figure 1C & D**) (p<0.05). Less prominent cells such as BEST4+ enterocytes and tuft cells were also detected, demonstrating the robustness of our annotation strategy (**Figure 1A & E**). Principal components extracted on cell type proportions demonstrated unique differences in cellular composition among the patient phenotype groups, while also showing patient-to-patient variabilities within the same phenotypic group (**Figure 1F**). These data pointed towards changes in cellular subtypes of TN_CD and Es_CD patients that might have diagnostic and alternative treatments potential.

**Figure 1.**
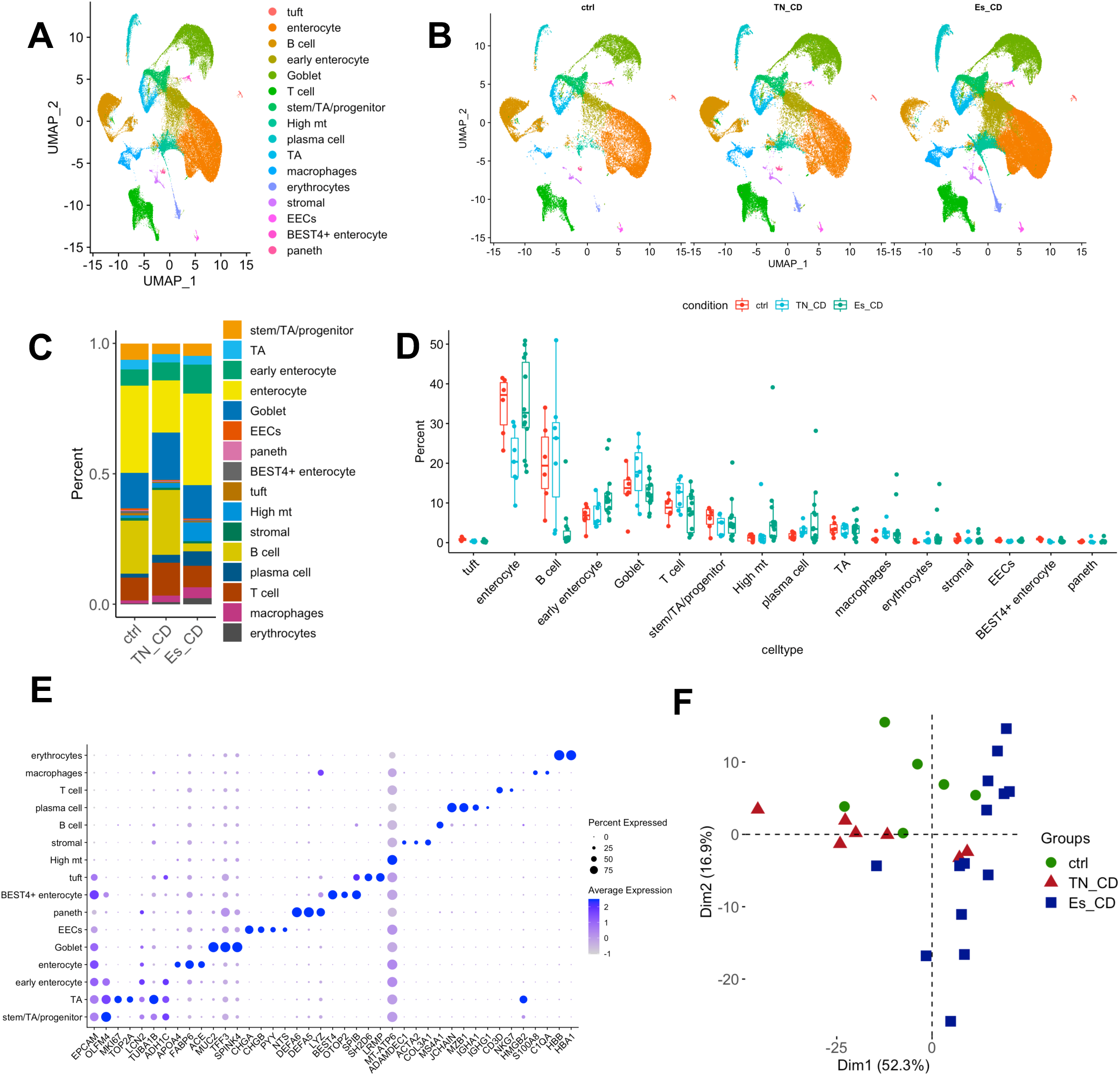
Human mucosal terminal ileal tissue heterogeneity: **A**. Uniform Manifold Approximation and Projection (UMAP) representation colored by cell types. **B**. Split UMAP based on CD phenotypes of ctrl, TN_CD and Es_CD. C. Bar plot showing the cell type proportion in each phenotype group. **D**. Box plot showing the cell type proportion differences among phenotype groups (green: ctrl; red: TN_CD; violet: Es_CD). **E**. Dot plot representing marker genes for each cell type. **F**. Principal component analysis (PCA) of cell type proportions, showing that the three phenotype groups can be separated based on cell type proportions.

### Increased secretory and reduced absorptive epithelial cells in treatment-naïve CD

To gain more insight into the cellular differences of Ileal mucosa that appeared to be taking place in the epithelium (n = 88,913; defined in **Figure 1**), deeper analysis was performed on the epithelial cells without the immune and stromal cells, thus increasing epithelial cell cluster granularity. A separate analysis without interference of immune and stromal cell genes allowed the pipeline to more clearly identify highly variable genes and principal components driven by the variability among epithelial cell types. In doing so, we identified low abundant M-cells (**Figure 2A**) and defined each cluster based on its unique gene expression patterns (**Figure 2B**). The 13 epithelial subtypes (**Supplementary Figure 2B**) included differentiated absorptive enterocytes, secretory goblet, EECs, pH-sensing absorptive BEST4+ enterocytes, chemosensory tuft cells, defensive secretory Paneth cells, low abundant M-cells, early enterocytes, proliferating cells, and four clusters of undifferentiated cells enriched for early enterocytes, along with TA, or stem/TA signatures (**Figure 2A and B**). The early enterocyte cluster was likely grouped based on cell cycle genes and precursor lineage genes for absorptive and secretory epithelial cell subtypes. The low abundant M-cells were missed in the first round of clustering analysis when immune and stromal cells were included.

Remarkably, the abundance of the M-cells, along with several other epithelial subtypes including tuft, goblet, TA, and BEST4+ cells appear to be altered in the mucosa of the TN_CD group compared to non-IBD ctrl (**Figure 2C**, p=0.035). Compared to the results from (**Figure 1)** when all the cell types were considered, here in the epithelial focused clustering analysis (**Figure 2**) the enterocytes in TN_CD patients were significantly reduced in proportions compared to ctrl (p=0.0047) and Es_CD (p=0.016). Both TN_CD and Es_CD patients had significantly decreased Best4+ enterocyte (p=0.0047 & 0.0087) and tuft cells (p=0.035 & 0.0066) compared to ctrl. TN_CD patients also had goblet cells in a higher abundance when (p=0.008 & 0.00012) compared to Es_CD and ctrl patients (**Figure 2C**). PCA analysis on epithelial cellular composition across patient phenotype groups (**Supplementary Figure 3**) showed that cell type changes were more pronounced in TN_CD compared to Es_CD and ctrl (**Figure 2D**). These data highlight differences in the epithelial cell subtype proportions related to disease and treatment, and points to cellular microenvironments of the epithelium that are altered during the early stages of untreated CD.

**Figure 2.**
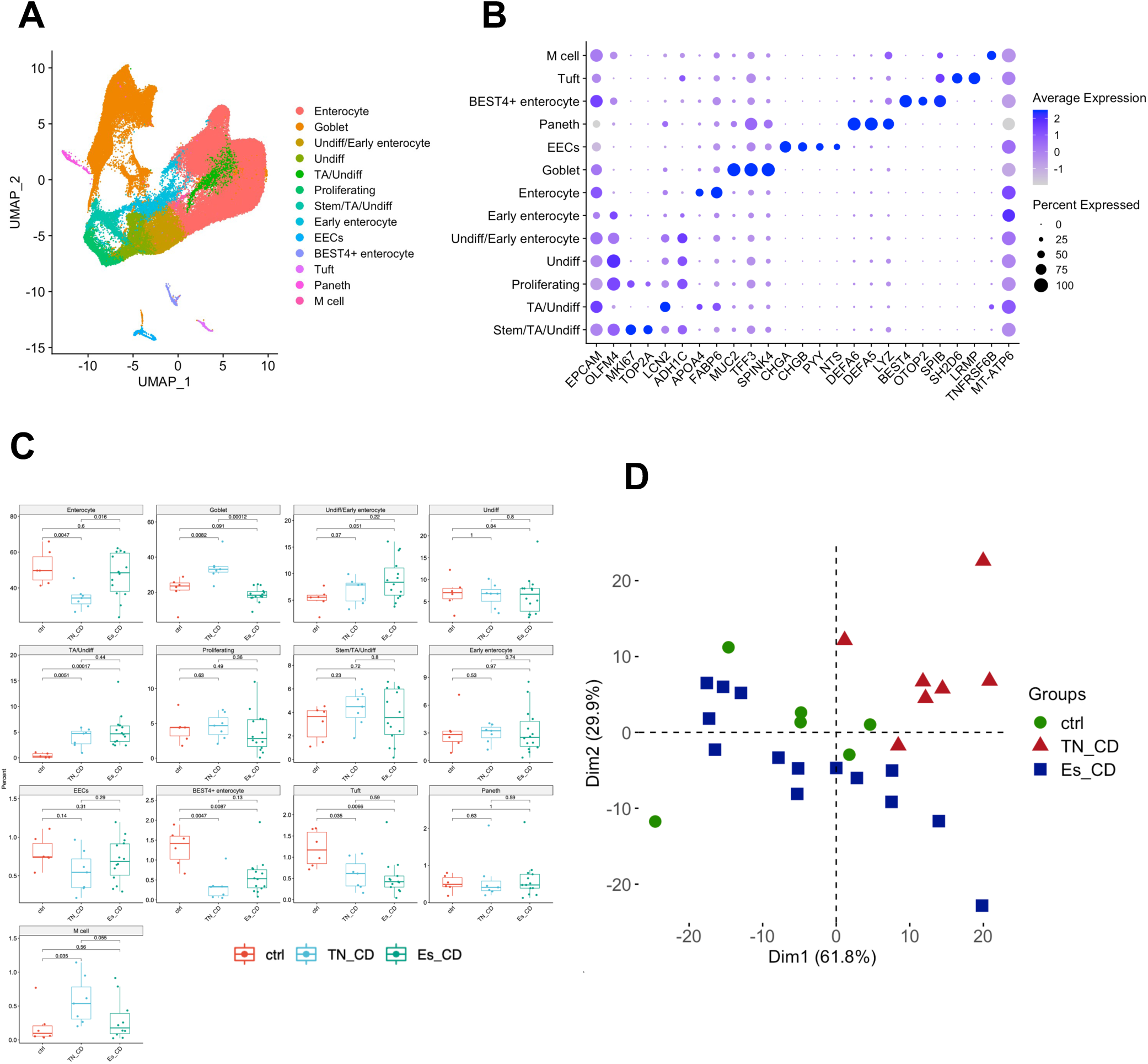
Epithelial sub-clustering: **A**. UMAP representation of epithelial cells, colored by 13 cell types. **B**. Dot plot representing marker genes for each epithelial cell type. **C**. Box plot showing the cell type proportion differences among the phenotype groups. **D**. PCA of cell type proportions, showing the variations of epithelial cell type compositions among the phenotype groups.

### Decreased B cells and heterogeneity in inflammatory macrophages in Es_CD

To gain insight into the cellular diversity and behavior of the immune compartment, we subclustered and analyzed the immune cells using a separate analysis without the epithelial and stromal cells (n = 43,648; defined in **Figure 1A**). In this analysis, we identified 9 immune cell subtypes (**Figure 3A**). The identified cell subtypes included B-cells, plasma cells, T-cells, erythrocytes, and a cluster showing mixed marker genes that we call unmatched cells (**Supplementary Figure 2C**). We were able to identify CD8+ T-cells and NK cells and distinguish inflammatory macrophages (inf. macrophage) from tissue resident macrophages (res.macrophages) (**Figure 3B**). Es_CD patients had significantly lower B-cell proportions compared to TN_CD & ctrl (**Figure 3C**) (p=0.0097 & 0.0064). Es_CD patients interestingly showed more unmatched cells when compared to TN_CD (p=0.031) & ctrl (p=0.00021) (**Figure 3C**). Inflammatory macrophages were found to be more abundant in Es_CD and TN_CD mucosa when compared to ctrl (p=5.2e10-5 & 0.022), but the increase in this type of macrophages were primarily contributed by a few patients, highlighting the patient-to-patient heterogeneity for some cell type proportions (**Supplementary Figure 4**). PCA analysis showed Es_CD group with strong differences in cell type composition compared to TN_CD and ctrl (**Figure 3D**), that was likely caused by the decrease in their B-cell populations. Taken together, we found a remarkable similarity in immune cell type abundance, suggesting the disease may be more related to the behavior of these cells than their proportions.

**Figure 3.**
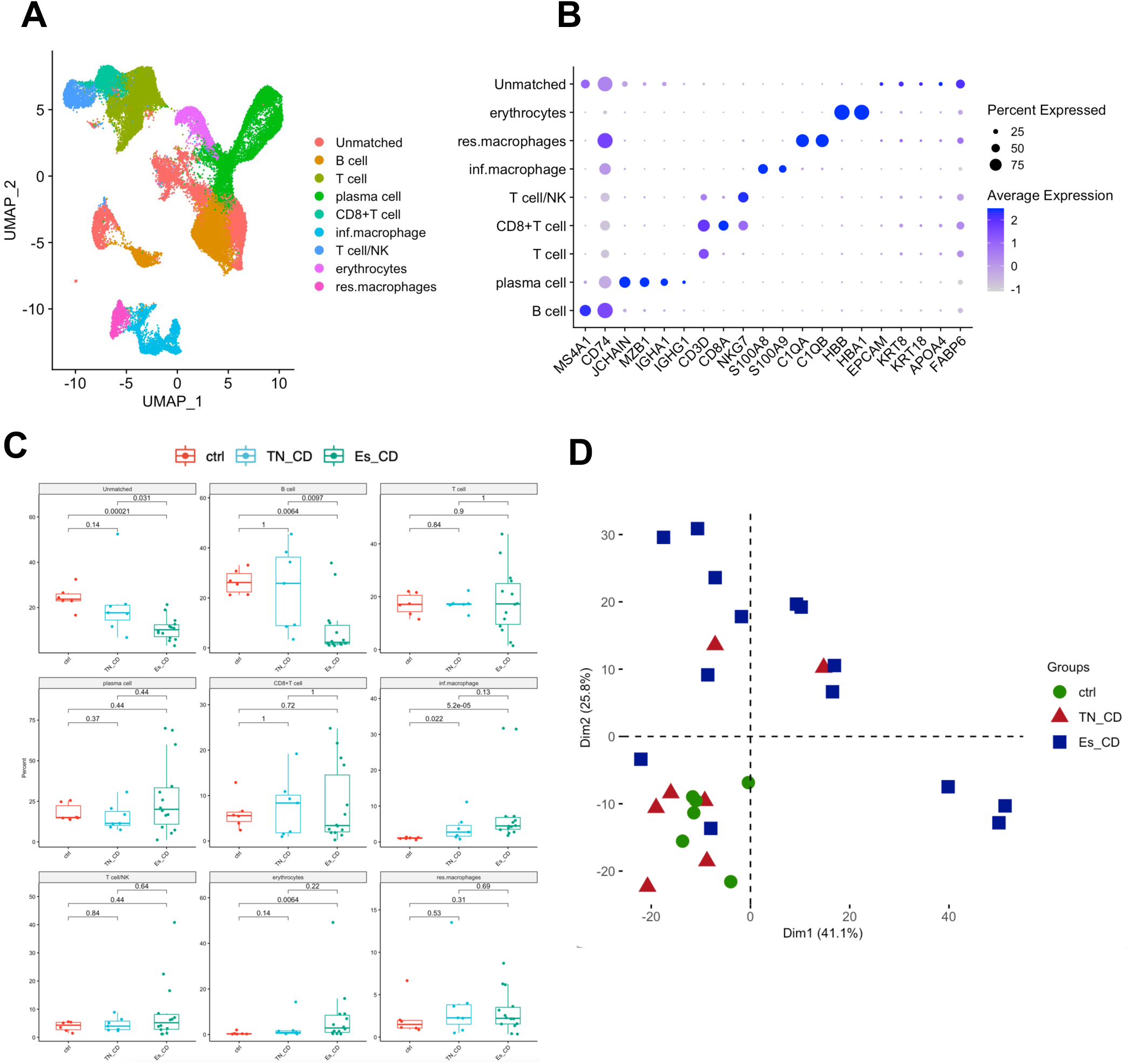
Immune sub-clustering: **A**. UMAP representation of immune cells, colored by 11 immune sub-clusters. **B**. Dot plot representing marker genes for each immune cell type. **C**. Box plot showing the cell type proportion differences among the phenotype groups. **D**. PCA of cell type proportions.

### Cell-type-specific expression changes in epithelial cell subtypes

To gain a closer insight into the behavior of the different mucosal cell types, we performed their cluster-specific differential gene expression (DGE) analysis. Here, we aim to identify cell-type-specific gene expression changes between the CD phenotype groups compared to controls in each epithelial cell type (**Supplementary Table 2**). This differential expression analysis found 18 genes highlighted in (**Figure 4A)**. Interestingly, both untreated (TN_CD) and treated (Es_CD) patients expressed more MHC class II protein complex binding genes, such as HLA-DP & HLA-DR, in most of the epithelial subtypes except EECs, Paneth and tuft cells when compared to controls, indicating a change in epithelial behavior as it begins to display more foreign antigen on its surface^28^. Likewise, both CD groups also showed MHC class I protein binding gene *HLA-B* expressed in almost all epithelial subtypes except EECs and Paneth when compared to controls. Additionally, this analysis showed that *CD74* transcript encoding a receptor linked to MHCII membrane transport^29^, inflammation and intestinal mucosal healing^30^, had increased transcript levels in most of the epithelial cell subtypes from Es_CD and TN_CD patients (**Figure 4A**) Furthermore, the M-cells showed expression of GWAS implicated IBD risk gene *CCL20* along with TA/undiff especially in TN_CD ^31^. When we analyzed the differences in cellular composition between remission and refractory mucosa, surprisingly, the epithelial cell type compositions between these groups appeared to be similar (**Figure 4B**). However, DGE analysis for each epithelial cell type between remission and refractory patients within the established CD group showed significant transcriptional differences (**Figure 4C**). It indicates that indeed in some instances it is not the composition of the cell types *per se* but more likely their activity, i.e gene transcription, that is contributing to the disease and progression. In fact, enterocytes and TA/undiff cell types from patients in remission have increased levels of *LCT, APOA1, APOA4*, and *RBP2* transcripts when compared to refractory. Goblet cells from the mucosa of patients in remission showed more expression of *TFF1* when compared to refractory. When the differentially expressed genes between CD and ctrl were used to perform Gene Ontology (GO) enrichment to identify biological processes involved in each sub type (**Figure 4D**), the predominate biological activity highlighted by this analysis indicated changes in antigen presentation, reactive oxygen production and other immune functions; processes that point to decreases in epithelial barrier function and an increase in exposure of the lamina propria to luminal contents.

**Figure 4.**
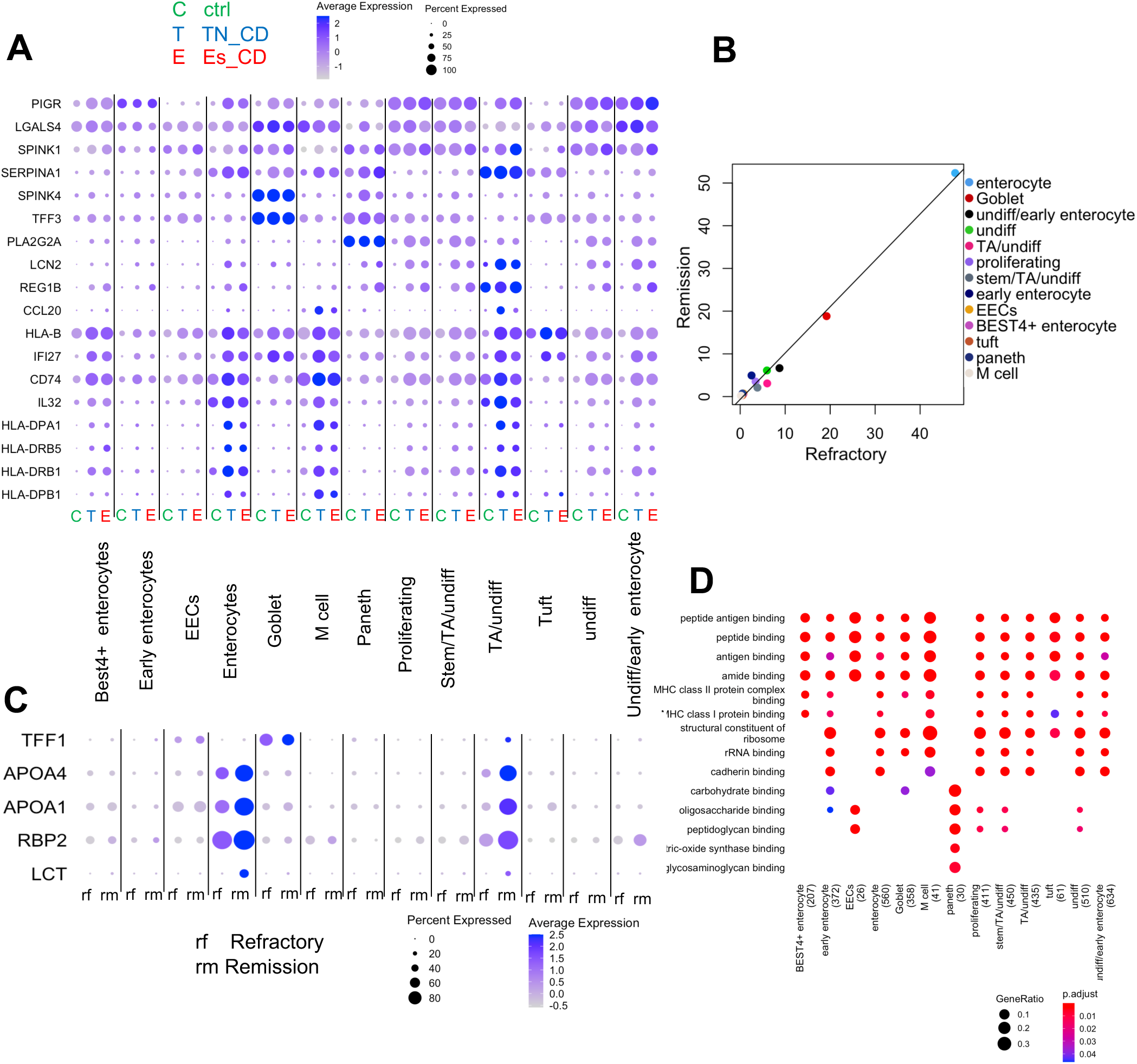
Epithelial cell type specific expression changes in Crohn’s Phenotypes: **A**. Dot plot showing differentially expressed genes in epithelial cell types between the phenotype groups. **B**. Scatter plot showing the epithelial cell type proportions between the remission and refractory are highly similar. **C**. Dot plot showing differentially expressed genes in epithelial cell types between remission and refractory. **D**. Dot plot showing GO enrichment of biological processes in epithelial cell types.

### Cell-type-specific expression changes in immune cell subtypes

The behavior of the immune compartment was also studied by DGE analysis for every immune cell subtype of each phenotype pair (**Supplementary Table 3**). Here, B-cells also showed changes in MHC class II expression like that of inflammatory macrophages, and plasma cells, while B- and T-cells showed higher expression of *BTG1* in TN_CD compared to non-IBD controls and Es_CD (**Figure 5A**). In the erythrocytes *ALAS2* expression was higher in TN_CD & Es_CD, while inflammatory macrophages showed higher expression of *HLA*-DR genes in controls and treatment naïve when compared to Es_CD. However, inf a few patients, inflammatory macrophages in Es_CD and TN_CD showed higher expression of inflammatory markers *S100A8, IL1B, SOD2* and *SAT1* that was not detected in resident macrophages. We also performed separate DE and GO analysis between the CD treatment groups, remission and refractory, and found that there were no significant proportional differences between immune subtypes in these two groups (**Figure 5B**) but we did find transcriptional differences (**Figure 5C**), further exemplifying the “behavior over abundance” hypothesis our data has helped to develop. For example, *HLA-DRB5* expression was increased in remission of inflammatory macrophages and refractory of resident macrophages, whereas *CCL5* expression was higher in T cell/NK in the remission group. GO analysis of the genes differentially expressed between CD and ctrl showed a strong metabolic signature along with changes in MHCII activity, along with numerous other processes related to immune function (**Figure 5D**). Taken together, a clear change in the activity of the immune compartment in the ileal mucosa is taking place during CD despite observing only minor changes in the relative abundance of different immune subtypes.

**Figure 5.**
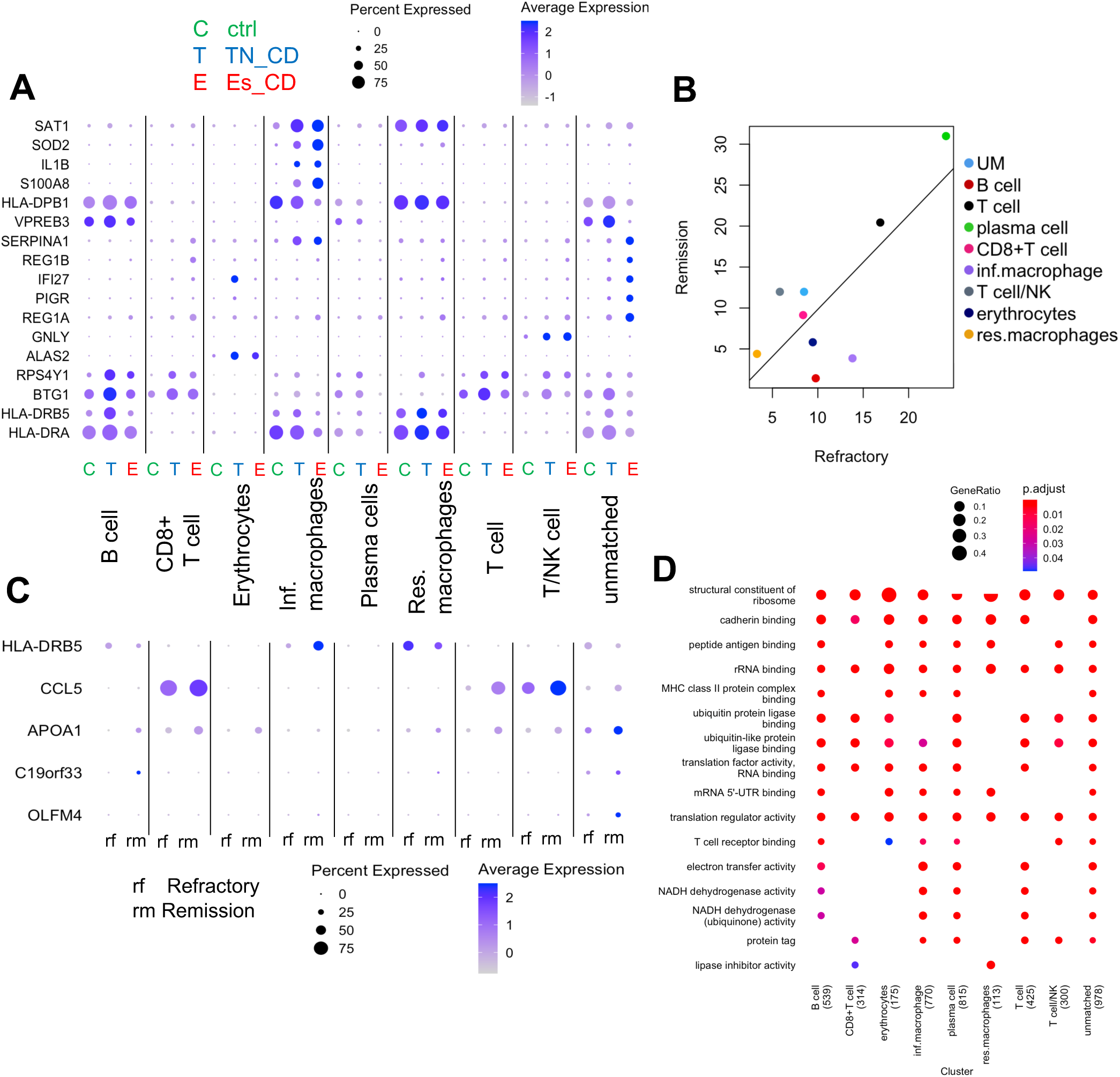
Immune cell type specific transcriptional changes in Crohn’s Phenotypes: **A**. Dot plot showing differentially expressed genes in immune cell types between the phenotype groups. **B**. Scatter plot showing the immune cell type proportions between remission and refractory are correlated. **C**. Dot plot showing the differentially expressed genes in immune cell types between remission and refractory. **D**. Dot plot showing GO enrichment of biological processes in immune cell types.

## DISCUSSION

Our primary goal in this investigation was to develop a robust and granular cell type atlas of the ileal mucosa during distinct stages of CD, including treatment-naïve and managed CD, along with refractory cases. We have relied upon scRNA-seq that has revolutionized numerous fronts in biology including cancer and cystic fibrosis^32^, to define the cell types and their activity by their transcriptomic signatures. We were driven by the hypothesis that changes in the cellular mucosa of CD would have profoundly different composition than that of non-IBD controls, particularly within the immune cell compartment, but we were surprised to find that although many of the patients had unique cellular abundance profiles, only a few disease-specific changes in cell type abundance stood out for CD as a whole, including changes in tuft and BEST4+, regardless of naïve, remission or refractory. What was striking in our data was the prominent rearrangement of the epithelial compartment in the TN_CD cohort, showing alterations in the abundance of enterocytes, M-, goblet, BEST4+, tuft, and TA cells. Furthermore, numerous transcriptional changes were detected between the phenotypic groups amongst similar cell types both in the immune and the epithelial compartments, which taken together clearly points to a dysfunctional epithelial barrier during CD and exposure of the underlying lamina propria to luminal contents that promote inflammatory activity in the surrounding mucosa.

We have identified both unique and patient-specific signals mapping to the epithelium of TN_CD. The alterations we found in epithelial abundance in TN_CD determined by scRNA-seq is supported by years of clinical and histological findings pointing toward degradation of the epithelium coincident with leaky barrier function and is a hallmark feature of newly diagnosed, particularly inflamed, CD^33, 34^. Loss of epithelial barrier leads to exposure of the lamina propria to an influx of luminal contents that initiate immune responses that we also detected in the form of MHCII expression, a process that can take place in both immune and non-immune cells^35, 36^. An earlier study also showed that intestinal M-cells with immune surveillance activity were induced during inflammation^37^ in colonic mucosa, supporting our observations in the ileum. Although we are not the first to show the importance of BEST4+ enterocytes^22, 24^ and tuft cells^38, 39^ to intestinal homeostasis and disease or changes in enterocytes and goblet cell levels in CD ^40^, we are for the first time, with the largest number of cells mapped from the terminal ileum in such a phenotypically diverse cohort to date, characterizing the global level of disruption in these cell subtypes and their activity within the TN_CD mucosa.

Although demonstrated by Suzuki et., al^41^ we did not find modifications in the stem cell compartment of the epithelium related to disease activities. However, we did observed significant increases in TA/undifferentiated cells along with *CDKN2A* senescence marker (data not shown) in the CD group compared to non-IBD controls, which might be related to the stem & TA/ undifferentiated cell activity shown by the studies of Wang et., al^42^ whereby senescence of transit amplifying cells were no longer able to proliferate into new enterocytes. In line with those findings, disruption of the proliferative capacity of the TA cells would likely result in loss of differentiated enterocyte abundance and contribute to epithelial erosion, villi disruption and decreased nutrition absorption.

The effects of therapy can be observed in our scRNA-seq data from the established cohort. Supported by results from Haberman el al who showed association of *APOA4* with better IBD outcome^43, 44^ in pediatric CD patients, our analysis showed the remission group had increased levels of *LCT, APOA1, APOA4*, and *RBP2* expression in enterocytes and TA/undifferentiated cell types when compared to treatment refractory patients. Other links to lipid metabolism, including those to *APOA1* and IBD have been established, with some therapeutic potential^44^, suggesting that disruption of the epithelial subtypes during CD impacts the absorbative and metabolic processes these cells normally perform, and may be linked to some of the nutritional difficiences and peripheral effects observed during CD. Moverover, alterations in goblet cell behavior was also observed between remission and refractory patients, a finding supported by Hensel et al^45^ that demonstrated increased mRNA expression levels of *TFF1, MUC1* and *HGF* in pediatric IBD during clinical remission. Thus, focusing on lipid metabolism and goblet cell function may be important targets in helping patients who are refractory to treatment. It should also be noted that where TN_CD had decreased epithelial numbers, the established patients, many of which are refracory to anti-TNF therapy, showed similar epithelial profiles to non-IBD controls indicating at least some form of mucosal healing after treatment.

Except for a few CD patients having increased levels of inflammatory macrophages in our cohort, along with an overall decrease in B-cell populations, no significant changes in the abundance of cells in the immune compartment were detected for CD patients as a whole. While changes in mucosal B-cell abundance was not surprising ^46, 47^, we were surprised to not find more cellular compositional changes in the immune compartment during CD, especially for those in the TN_CD group. For example, the absence of macrophages having significantly higher expression of inflammatory markers such as *S100A8*^*48*^, *IL1B*^*49*^, *SOD2*^*50*^ and *SAT1* in most of our diseased patients suggested alternative mechanisms contributing to disease than just changes in cell subtype abundance or an influx of inflammatory cells. This has led us to consider a new hypothesis that it in some instances it is the cellular activity involving gene transcription, protein function and cellular crosstalk that is playing a larger role in disease onset and progression than it is a change in cellular composition.

In support of this new hypothesis, our analysis showed higher levels of a well-studied IBD chemokine *CCL20* in M-cells of TN_CD and found increases in *CCL5* production in T- and NK cells. Recent work from our group using patient derived intestinal organoids has begun to show the importance of the epithelium in CD and its role in producing soluble molecules that can impact the behavior of surrounding cell types including those in the immune compartment ^7, 51^. Studies of Kaser et., al^52^ and Smillie et. al.,^31^ showed *CCR6* receptor and its ligand *CCL20* were upregulated in both CD and ulcerative colitis (UC) in 114 patients and is a GWAS implicated risk gene^31^ for UC. Furthermore, our analysis showed that epithelial sub clusters such as enterocytes, M-cells and TA/undifferentiated cells had significant increases in *HLA-DP, HLA-DR* and *HLA-B* in CD patients with a fold change greater than 2. This pattern of expression by the epithelium is directly involved in antigen presentation pathways and processing^53^, indicating these cells have been stimulated by microbes and other luminal contents allowed to cross the damaged epithelial barrier and are displaying foreign antigens to prompt an immune response. Alternatively, changes in the microbiome of some of these patients may also precipitate alterations in MHCII expression. Nonetheless, our single cell data has enabled us to further substantiate the idea that it could be the crosstalk across cell subtypes and the modulation of cellular behavior that is more important to disease in some cases than changes in cellular abundance.

In summary, to better understand the differences in cell type and gene expression profiles between treatment-naïve patients and established patients with therapy, we used the revolutionary technology of scRNA-seq and subsequent bioinformatics to analyze 27 patients, generating single cell transcriptomic data for ~130K cells that were annotated into epithelial^22, 24,38^, immune^9^ and stromal^54^ cell subtypes to reveal an overwhelming change in the mucosal epithelium of TN_CD along with numerous changes in immune and epithelial activity in distinct disease states. Our analysis showed cell type abundance and transcriptional heterogeneity that was unique to the individual and in some cases the disease and gives new direction in designing personalized therapies and possible explanations for treatment failure. Our study was not without limitations, however, in that we only captured a subset of the genes expressed by these cells, the spatial organization of these cells were lost upon tissue dissociation, and we did not look at protein levels or surface expression on the cells. Despite these limitations we have shown an important role of epithelial dysfunction during the early stages of CD, and future studies using spatial transcriptomics on mucosal tissue sections along with CITE-seq that couples cells surface protein detection with sequencing will be used to overcome these limitations and further refine and granulate our analysis.

## Data Availability

All data produced in the present study are available upon reasonable request to the authors

## Abbreviations

(CD): Crohn’s Disease
(Es_CD): Established CD
(GEMs): Gel Bead in Emulsion
(GO): Gene Ontology
(GWAS): Genome wide associations Studies
(CyTOF): Mass Cytometry
(M cells): Microfold cells
(RPCA): Reciprocal Principal Component Analysis
(SESCD): Simple Endoscopic Score for Crohn Disease
(scRNA-seq): Single cell RNA sequencing
(TN_CD): Treatment Naïve CD
(UMAP): Uniform Manifold Approximation and Projection

## Data availability

The scRNA-seq data for the 27 patient’s terminal ileal biopsies included in this study has been deposited in Gene Expression Omnibus (GEO) and is accessible through GEO accession GSE202052

## Acknowledgement

We are grateful to Murugadas Anbazhagan and Greg Gibson for their support and helpful comments. We acknowledge the Genomic Cores at Yerkes Non-Human Primate Research Center at Emory University (NIH P51 OD011132; NIH S10 OD026799), and the Parker H. Petit Institute for Bioengineering and Bioscience at the Georgia Institute of Technology for the single-cell library sequences.

