## Supplementary materials for "Assessing Cellular and Transcriptional diversity of IIeal Mucosa amongst Treatment Naïve and Treated Crohn’s disease"

**Cellular ileal composition and their behavior during Crohn’s disease**

Sushma Chowdary Maddipatla­­^1^, Vasantha L Kolachala^1^, Suresh Venkateswaran^1^, Anne F Dodd­­^1^, Ranjit Singh Pelia^1^, Duke Geem^1^, Hong Yin^2^, Yutong Sun^3^, Congmin Xu^4^, Angela Mo^5^, Astrid Kosters^6^, Junkai Yang^6^, Jason D. Matthews^1^, Eliver Ghosn^7^, Subra Kugathasan^1,8,9, $, *^ and Peng Qiu^4, $, *^

**Supplementary Materials**

**Supplementary Figure Legends**

Supplementary Figure 1. Quality Control: Density plots showing the number of UMIs per cell (A), number of genes detected per cell (B) for the three CD phenotype groups and for each cell type (C), where pink represents ctrl, green represents TN, and blue represents Es. Performance of data integration and batch correction is shown in UMAPs colored by phenotype groups (D & E), showing the overlap of cell types among the phenotype groups.

Supplementary Figure 2: Validation of tissue compartments based on marker genes: Scatter plots of cell clusters along three axes, where each axis represents the average expression of epithelial/immune/stromal marker genes in each cell cluster. Each dot in the scatter plots represents one cell cluster. The fact that majority of the cell clusters locate close to the axes validated tissue compartment of the cell clusters.

Supplementary Figure 3: Epithelial cell type proportions: Bar plot showing the epithelial cell type proportions for every sample.

Supplementary Figure 4: Immune sub-type proportions: Bar plot showing the immune cell type proportions for every sample.

Supplementary Figure 5: Phylogenetic tree: Phylogenetic tree of the epithelial (A) and immune (B) cell types respectively.

**Supplementary Table Legends**

Supplementary Table 1. Patient demographics: Total number of controls, Treatment-naïve and established CD patient’s terminal ileal biopsies utilized for single cell RNA sequencing including breakdown on their clinical information.

Supplementary Table 2. Differentially expressed genes in epithelial cell types: Epithelial cells from first round of clustering were subjected to re-clustering. Table includes differentially expressed genes between CD phenotypes for every epithelial cell type. ‘pct.1’ refers to percent of cells in that CD phenotype for a given epithelial cell type expressed the gene. ‘pct.2’ refers to percent of cells in other compared CD phenotype for a given epithelial cell type expressed the gene. ‘avg_logFC’ refers to average log fold change of level of gene expression in CD phenotype compared to other for a given epithelial cell type. P values of both adjusted and unadjusted are showed along with these.

Supplementary Table 3. Differentially expressed genes in immune cell types between the conditions: Immune cells were sub-clustered, and the table includes differentially expressed genes for every immune cell type between the CD phenotypes. ‘pct.1’ indicates to percent of cells expressing that gene in that CD phenotype for a given Immune cell type. ‘pct.2’ refers to percent of cells expressing that gene in other compared CD phenotype for a given immune cell type. ‘avg_logFC’ refers to average log fold change of gene expression levels between the CD phenotype for a given immune cell type. P values of both adjusted and unadjusted are showed along with these.
