## Supplementary figures and images for "Assessing Cellular and Transcriptional diversity of IIeal Mucosa amongst Treatment Naïve and Treated Crohn’s disease"

### Supplementary figure 1

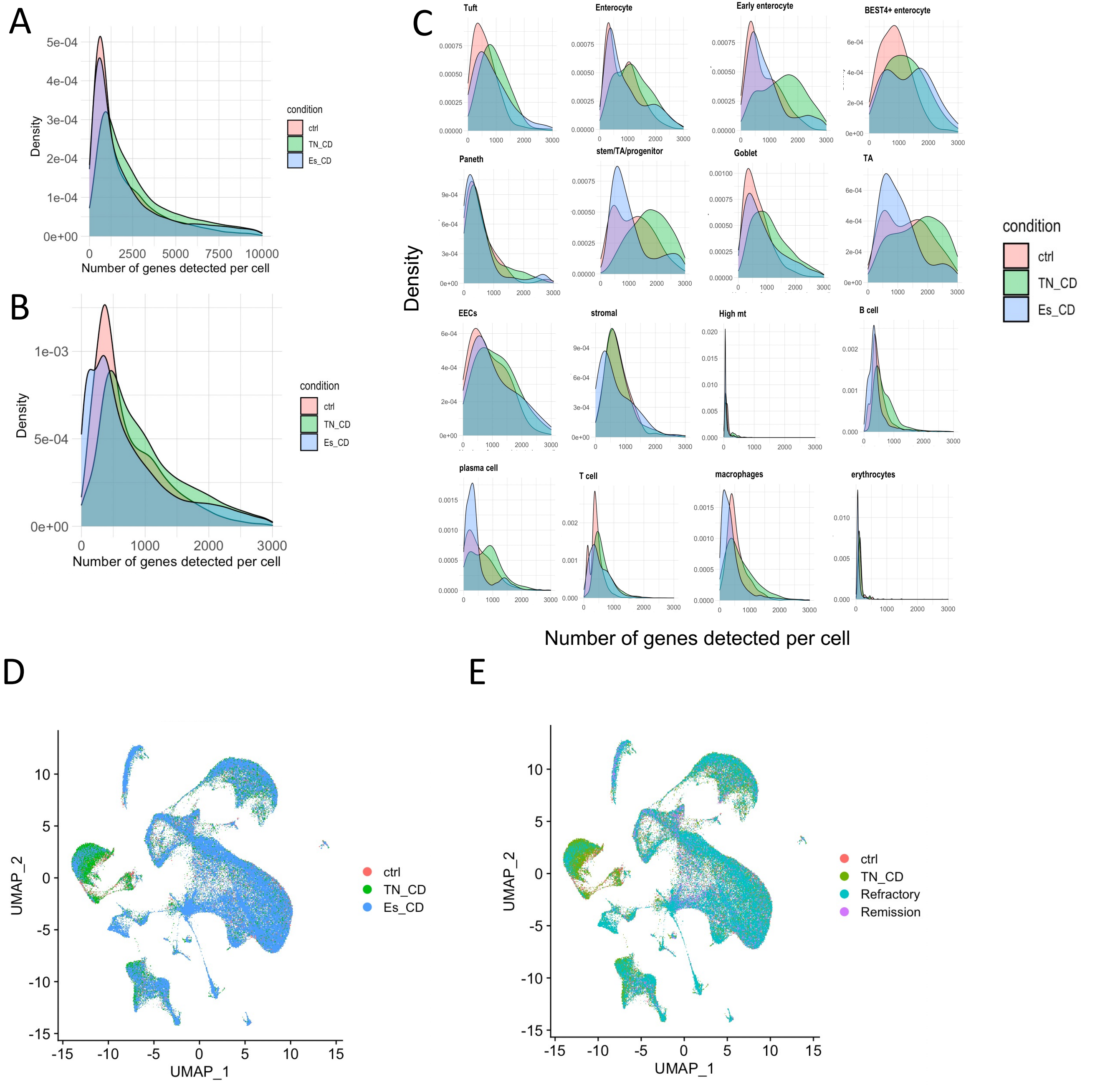

### Supplementary figure 2

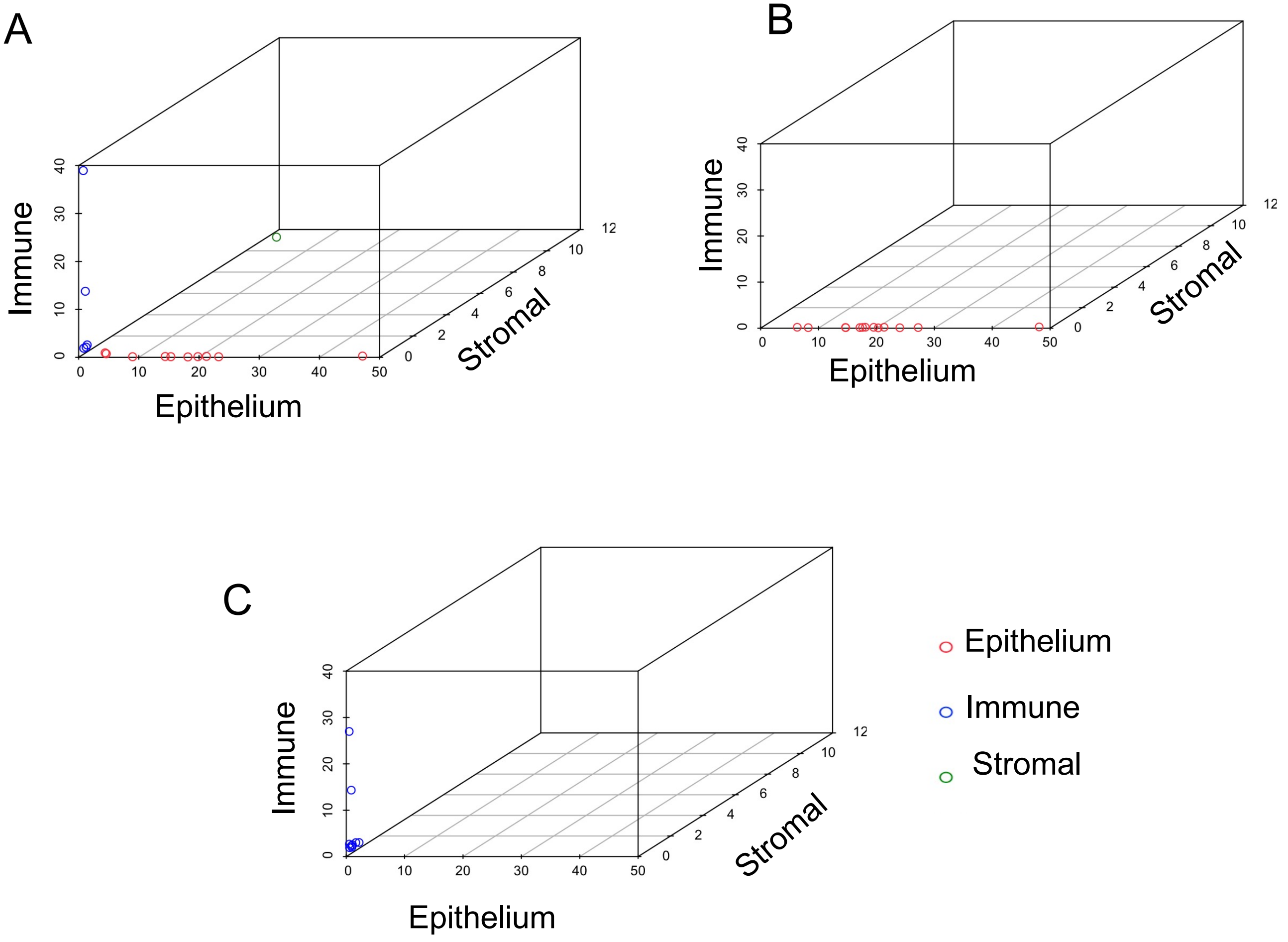

### Supplementary figure 3

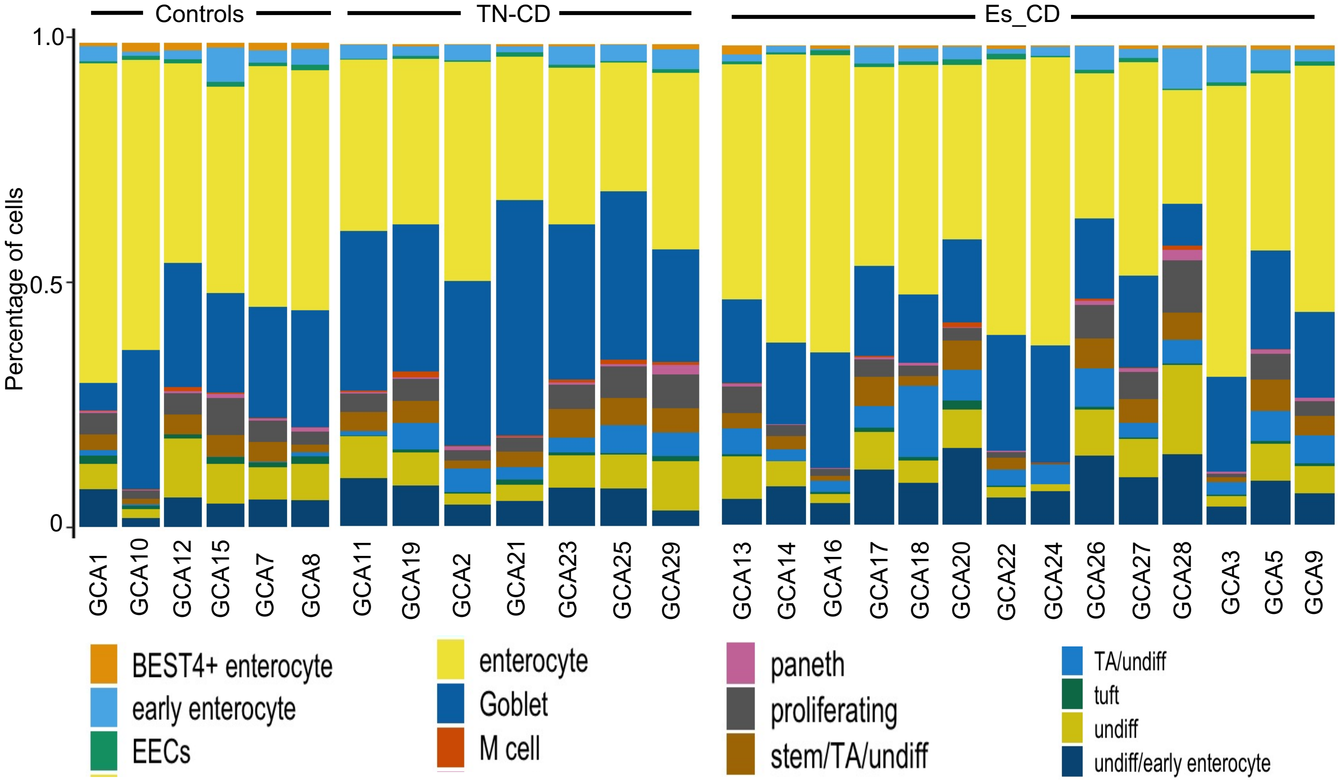

### Supplementary figure 4

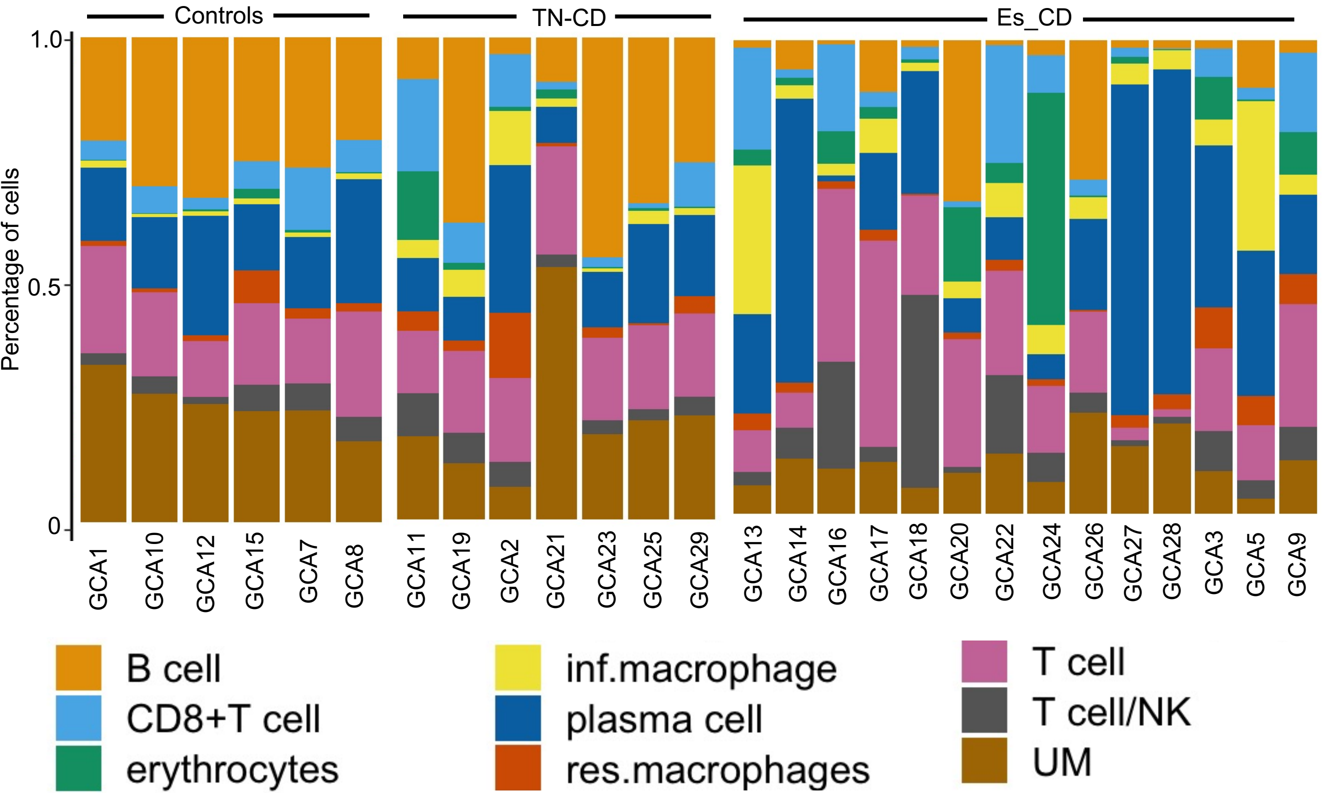

### Supplementary figure 5

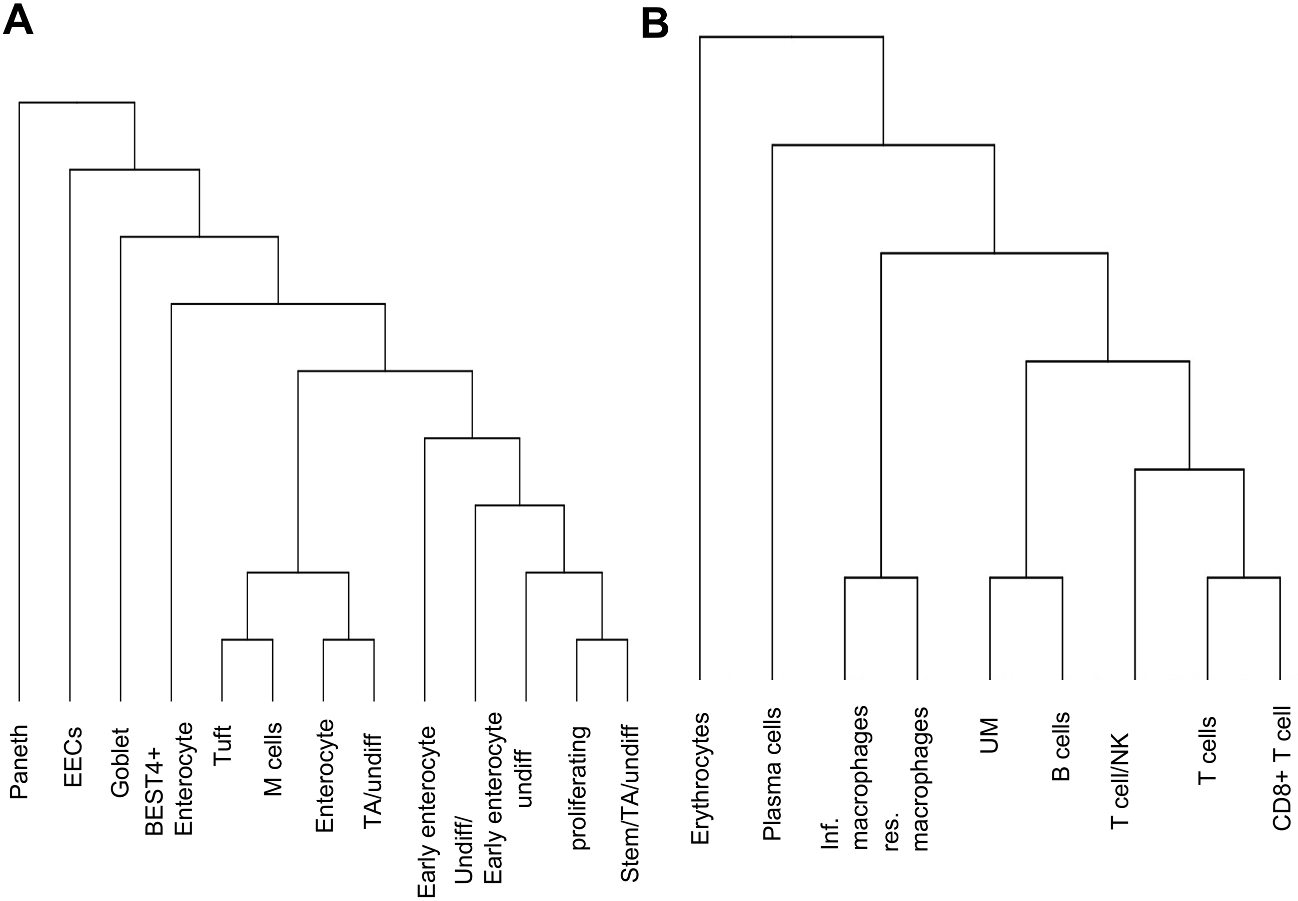
